# Segregation analysis of 17,425 population-based breast cancer families: evidence for genetic susceptibility and risk prediction

**DOI:** 10.1101/2022.05.24.22275555

**Authors:** Shuai Li, Robert J. MacInnis, Andrew Lee, Tu Nguyen-Dumont, Leila Dorling, Sara Carvalho, Gillian S. Dite, Mitul Shah, Craig Luccarini, Qin Wang, Roger L. Milne, Mark A. Jenkins, Graham G. Giles, Alison M. Dunning, Paul D.P. Pharoah, Melissa C. Southey, Douglas F. Easton, John L. Hopper, Antonis C. Antoniou

## Abstract

Rare pathogenic variants in known breast cancer susceptibility genes and known common susceptibility variants do not fully explain the familial aggregation of breast cancer. To investigate plausible genetic models for the residual familial aggregation, we studied 17,425 families ascertained through population-based probands, 86% of whom were screened for pathogenic variants in *BRCA1, BRCA2, PALB2, CHEK2, ATM* and *TP53* using gene-panel sequencing. We conducted complex segregation analyses and fitted genetic models in which breast cancer incidence depended on the effects of pathogenic variants in known susceptibility genes and other unidentified major genes, and a normally distributed polygenic component. The proportion of familial variance explained by *BRCA1, BRCA2, PALB2, CHEK2, ATM* and *TP53* was 46% at age 20-29 years and decreased steadily with age thereafter. After allowing for these genes, the best fitting model for the residual familial variance included a recessively inherited risk component with a combined genotype frequency of 1.7% (95% CI: 0.3-5.4%) and a penetrance to age 80 years of 69% (95% CI: 38-95%) for homozygotes, and a polygenic variance of 1.27 (95% CI: 0.94-1.65) which did not vary with age. The proportion of the residual familial variance explained by the recessive risk component was 40% at age 20-29 years and decreased with age thereafter. The model predicted age-specific familial relative risks consistent with those observed by large epidemiological studies. The findings have implications for strategies to identify new breast cancer susceptibility genes and improve breast cancer risk prediction, especially at a young age.

## INTRODUCTION

There is a substantial familial aggregation of breast cancer. The familial relative risk (FRR) of breast cancer for having an affected first-degree relative is on average about 1.8 but varies substantially by the age at cancer diagnosis of the relative(s), the number of affected relatives, and the age of the consultant^1^.

High-risk pathogenic variants (PVs) in the currently known breast cancer susceptibility genes *BRCA1, BRCA2* and *PALB2* and intermediate-risk PVs in genes such as *CHEK2* and *ATM*^2-10^ on average explain 20-25% of the familial aggregation of breast cancer, and much more at younger ages. A further on average ∼20% of the familial aggregation is accounted for by a polygenic risk score (PRS) based on 313 common genetic variants identified by genome-wide association studies (GWAS)^11^. PVs in other genes including *BARD1, RAD51C, RAD51D, TP53, PTEN* and *NF1* are also associated with breast cancer risk, but to a lesser extent and account for a small proportion of the familial aggregation^2^. Consequently, a large proportion of the familial aggregation remains unexplained.

Segregation analyses have been used to develop pedigree-based statistical models of breast cancer susceptibility and predict breast cancer risk based on family cancer history, genotype and other factors^**12-15**^. For example, the Breast and Ovarian Analysis of Disease Incidence and Carrier Estimation Algorithm (BOADICEA) for estimating a woman’s future risk of developing breast cancer^16-20^, was originally developed using data on 2,785 families and considered breast cancer familial risk to be determined by the joint effects of *BRCA1* and *BRCA2* PVs and a polygenic component representing the combined multiplicative effects of a large number of unknown genetic variants each making a small contribution to the variation in risk^12,14,17^. The model was subsequently extended to incorporate PVs in *PALB2, CHEK2* and *ATM*, polygenic risk scores, lifestyle- and hormone-related risk factors and mammographic density^20^, and more recently PVs in *BARD1, RAD51C* and *RAD51D*^21^.

It was not previously possible to fit segregation analysis models that included the joint effects of all known breast cancer susceptibility genes simultaneously due to the lack of data on PVs in some genes. Another challenge has been small sample sizes that limit the statistical power to distinguish different inheritance models. In this study, we conducted the analysis of this kind using data from families ascertained through large population-based series of affected and unaffected probands for which data on PVs in *BRCA1, BRCA2, PALB2, CHEK2, ATM* and *TP53* were available. Our objectives were: (i) to estimate key genetic model parameters simultaneously to further improve the accuracy of existing genetic models; and (ii) to investigate the genetic models of inheritance that best explain the familial aggregation of breast cancer not accounted for by the known breast cancer susceptibility genes and polygenic factors.

## MATERIALS AND METHODS

### Study sample

The sample included 17,425 three-generation families ascertained via population-based sampling of breast cancer probands from two studies: 2,712 families from the population-based case-control-family study within the Australian Breast Cancer Family Registry (ABCFR) and 14,713 families from the prospective Studies of Epidemiology and Risk Factors in Cancer Heredity (SEARCH) study in the UK.

ABCFR^22-24^ includes: (i) 1,644 case families, ascertained independently of their family cancer history through a sample of adult women living in the metropolitan areas of Melbourne and Sydney who were diagnosed between 1992 and 1999 (baseline) with a histologically confirmed first primary breast cancer of (case probands) before age 70 years; and (ii) 1,068 control families ascertained through unaffected adult women (control probands) who were sampled at the same time using the Australian electoral rolls and frequency matched to case probands by age. Of these, 858 case families with a proband diagnosed with breast cancer before age 40 years were included in a previous segregation analysis^13^. Case and control probands gave a blood sample and completed the same risk factor questionnaire and family cancer history questionnaire involving the construction of a pedigree covering all known first- and second-degree adult relatives. In addition, each proband was asked to obtain permission from first- and second-degree relatives for their participation, which involved giving a blood sample, completing the same risk factor questionnaire, and providing additional information to complement the pedigree and family history information collected from the proband. This analysis used demographic data and breast and ovarian cancer diagnoses of the probands and all their adult female first- and second-degree relatives identified at baseline. ABCFR was designed to be enriched for cases diagnosed with breast cancer at younger ages: 55% of case probands and 52% of control probands included in this analysis were younger than 40 years at diagnosis and recruitment, respectively. Of the probands, 92% reported having white ethnicity.

SEARCH ^25^ ascertained families through adult women diagnosed with breast cancer, identified through the Eastern Cancer Registration and Information Centre. Eligible women were those diagnosed between 1991 and 1996 before 55 years of age and recruited between 1996 and 2002 (prevalent cases), together with women diagnosed between 1996 and 2011 before age 70 years (incident cases). Probands were invited to provide a blood sample and complete an epidemiological questionnaire, including family cancer history in all first-degree relatives and grandparents. This analysis used the demographic data and breast and ovarian cancer diagnoses of the probands and all their adult female first-degree relatives and grandmothers collected at baseline. A total of 1,484 families with a proband diagnosed before age 55 years between 1991 and 1996 were included in previous segregation analyses^12,14^. Of the probands included in this analysis, 94% were older than 40 years at diagnosis. Approximately three-quarters of the probands reported their ethnicity; 99% of which had white European ancestry.

ABCFR was approved by the Human Research Ethics Committee of the University of Melbourne. SEARCH was approved by the National Research Ethics Service Committee East of England— Cambridge South. All participants provided written consent.

### PVs in breast cancer susceptibility genes

We studied six major cancer susceptibility genes: *BRCA1, BRCA2, PALB2, CHEK2, ATM* and *TP53*. PVs included predicted protein-truncating variants (PTVs), and an additional subset of rare missense variants (population frequency <0.001) for *BRCA1, BRCA2* and *TP53*. The risks associated with this subset of missense variants have been shown to be similar to those associated with PTVs^2^, and these were aggregated with the PTVs in the analysis.

In ABCFR, PVs were identified by gene-panel testing for 2,305 probands (85% of all probands) and 770 relatives, and by other tests conducted before gene-panel testing was widely available (Table S1) for 2,317 probands and 1,765 relatives; 2,244 probands and 611 relatives tested by both regimes. In total, 88% of families were tested; see Southey et al^26^ for more details about the gene-panel testing and pathogenicity definition. In SEARCH, PVs were identified by gene-panel testing for 12,654 (86% of all probands) probands. No relatives were screened; see Dorling et al^2^ for more details about the testing and pathogenicity definition.

### Statistical methods

We conducted complex segregation analysis using the pedigree analysis software MENDEL following a similar approach to that used in previous studies^12-14^. Each woman was considered to be at risk of breast and ovarian cancer from birth until breast or ovarian cancer diagnosis, baseline interview, death, or age 80 years, whichever occurred first. The incidences of the two cancers were modelled simultaneously and assumed to be independent, conditional on genotype. For a woman *i* in birth cohort *k*, from country *c* and at age *t*, the incidence of cancer *s, λ*_*i*_*(t, k, c, s)*, was assumed to depend on the genetic factors according to the following model:

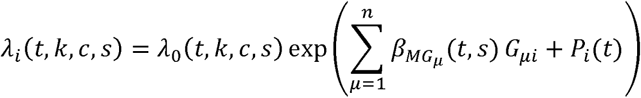

where *λ*_*0*_*(t, k, c, s)* is the baseline incidence, 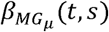 is the age-specific log-relative risk (log-RR) for a major gene that was assumed to be the same across countries and birth cohorts, *n* is the number of major genotypes, and *G*_*μi*_ is an indicator variable which takes the value of 1 if the woman has major genotype *μ* and 0 otherwise. *P*_*i*_*(t)* is the breast cancer age-specific polygenic component that was assumed to be normally distributed with zero mean and variance σ_P_^2^(t), the same across countries and birth cohorts, representing the multiplicative effects of a large number of variants each associated with a small increment in breast cancer risk. No polygenic component was included for ovarian cancer. The polygenic component was approximated by the hypergeometric polygenic model (HPM)^27,28^ as

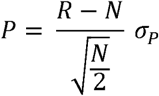

where *R* has a binomial distribution (2N, ½) and *N*, the number of loci used in the HPM, was 3. We firstly fitted a model with a polygenic component only, without any major genes. The polygenic variance σ_P_^2^(t) from this model reflects the total breast cancer familial variance under the polygenic susceptibility model^29^. Note that while σ_P_^2^(t) is termed the polygenic variance, it also captures the effects of non-genetic factors contributing to the risks for relatives being correlated, and whose existence is suggested by analysis of twin pairs in the Nordic Twin Study^30^. Given that breast cancer FRR decreases with age^1^, we allowed σ_P_^2^(t) to decrease linearly with age.

We then fitted a model equivalent to the BOADICEA (Version 4), which included the effects of PVs in *BRCA1, BRCA2, PALB2, CHEK2* and *ATM*, as well as the polygenic component. We sequentially extended this model to include the effects of PVs in *TP53*, and a hypothetical major gene for which we investigated different models of inheritance (dominant, recessive, general). The decrease in σ _p_^2^(t) as major genes were included was used to express the proportions of breast cancer familial variance explained by PVs in those genes.

To reduce computational time, the major gene component was fitted using a single locus comprising *m +* 1 alleles, where *m* is the number of major genes considered in the model. That is, we assumed separate risk alleles representing the presence of a PV in *BRCA1, BRCA2, PALB2, CHEK2, ATM, TP53* and the hypothetical gene, respectively, and a normal allele. We assumed a dominant inheritance for the risk alleles in the order of the seven major genes above; therefore, there was a total of nine possible major genotypes *μ: BRCA1* PV carriers, *BRCA2* PV carriers, *PALB2* PV carriers, *CHEK2* PV carriers, *ATM* PV carriers, *TP53* PV carriers, the hypothetical gene PV homozygotes, the hypothetical gene PV heterozygotes, and non-PV carriers. This simplification is unlikely to affect results because carriers of PVs in more than one major gene are very rare and would not contribute materially to the analysis; from our data, 0.1% of the women were such carriers (Table 1). To compute the baseline incidence *λ*_*0*_*(t, k, c, s)*, we used the method previously described^12^ to constrain the overall incidence across all genotypes to agree with the UK and Australian birth cohort-specific smoothed population incidences used in the BOADICEA^18^.

**Table 1.** Number of probands by study and the diagnosis age and PV status of the proband ^a^.

| Age group (years) | Total | *BRCA1* PV carriers | *BRCA2* PV carriers | *PALB2* PV carriers | *CHEK2* PV carriers | *ATM* PV carriers | *TP53* PV carriers | Non-PV carriers | Untested |
| --- | --- | --- | --- | --- | --- | --- | --- | --- | --- |
| **ABCFR case families** |  |  |  |  |  |  |  |  |  |
| <30 | 88 | 7 | 6 | 1 | 2 | 2 | 3 | 60 | 7 |
| 30-39 | 810 | 43 | 27 | 1 | 11 | 5 | 5 | 628 | 91 |
| 40-49 | 368 | 15 | 5 | 4 | 6 | 3 | 0 | 313 | 23 |
| 50-59 | 346 | 2 | 6 | 5 | 2 | 3 | 3 | 315 | 10 |
| 60-69 | 32 | 0 | 0 | 0 | 0 | 1 | 1 | 27 | 3 |
| Total | 1644 | 67 | 44 | 11 | 21 | 14 | 12 | 1343 | 134 |
| **ABCFR control families** |  |  |  |  |  |  |  |  |  |
| <30 | 86 | 0 | 1 | 0 | 1 | 3 | 0 | 55 | 26 |
| 30-39 | 468 | 2 | 3 | 1 | 0 | 1 | 0 | 366 | 96 |
| 40-49 | 240 | 2 | 1 | 0 | 2 | 0 | 0 | 206 | 39 |
| 50-59 | 218 | 0 | 1 | 0 | 1 | 2 | 1 | 179 | 35 |
| 60-69 | 56 | 0 | 0 | 0 | 0 | 0 | 0 | 42 | 14 |
| Total | 1068 | 4 | 6 | 1 | 4 | 6 | 1 | 848 | 210 |
| **SEARCH** |  |  |  |  |  |  |  |  |  |
| <30 | 60 | 4 | 2 | 1 | 3 | 0 | 1 | 35 | 15 |
| 30-39 | 878 | 19 | 29 | 1 | 15 | 1 | 5 | 614 | 194 |
| 40-49 | 3519 | 40 | 72 | 23 | 53 | 21 | 6 | 2665 | 642 |
| 50-59 | 5694 | 27 | 79 | 39 | 83 | 37 | 1 | 4648 | 782 |
| 60-69 | 4558 | 10 | 41 | 18 | 49 | 31 | 1 | 3983 | 426 |
| 70-79 | 4 | 0 | 0 | 0 | 0 | 0 | 0 | 4 | 0 |
| Total | 14713 | 100 | 223 | 82 | 203 | 90 | 14 | 11949 | 2059 |

<sup>a</sup> 10 Probands had a PV in two genes: ABCFR case families – one proband (30-39 years) in *BRCA1* and *BRCA2*, and one proband (40-49 years) in *BRCA1* and *CHEK2*; ABCFR control families – one proband (30-39 years) in *BRCA2* and *ATM*; SEARCH – two probands (40-49 years, 60-69 years) in *BRCA1* and *CHEK2*, one proband (50-59 years) in *BRCA2* and *PALB2*, and four probands (one <30 years, two 40-49 years, and one 50-59 years) in *BRCA2* and *CHEK2*

The age-specific breast and ovarian cancer RRs 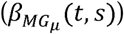 for *BRCA1* and *BRCA2* PV carriers, and breast cancer RRs for *PALB2, CHEK2* and *ATM* PV carriers, were fixed at estimates from previous studies^5,7,9,17^. Ovarian cancer RRs were assumed to be 1 for *PALB2, CHEK2* and *ATM* PV carriers, i.e., ovarian cancer incidence depended on *BRCA1* and *BRCA2* PVs only. The risk allele frequencies for all major genes, the age-specific RRs for *TP53* PV carriers and hypothetical gene PV carriers, and the age-specific polygenic variance σ_P_^2^(t) were estimated. The variances of the parameter estimates were obtained by inverting the observed information matrix. To allow for the restricted ranges of the parameter values and provide estimates likely to be more nearly normally distributed, we used transformed values for the parameters in the model: allele frequencies were logit transformed, RRs were log transformed, and σ_P_^2^(t) was square-root transformed. We compared nested models using the likelihood ratio test, and non-nested models using the Akaike Information Criterion. All statistical tests were two-sided, and results with a *P*<0.05 were treated as statistically significant.

Since not all PVs would be detected by the test methods used, we included a test sensitivity parameter defined as the probability of detecting a PV if one exists. In ABCFR, the test sensitivity was the weighted sum of the sensitivities of the test methods used (Table S1), with weights being the proportional lengths of the exons screened. The test sensitivity was assumed to be 100% for the relatives who had been only tested for their probands’ PV. For ABCFR probands, the average test sensitivity ranged from 88% to 90% across all genes. For SEARCH probands, the test sensitivity was assumed to be 90% for all genes. We also conducted a sensitivity analysis by assuming the test sensitivity to be 80% for all genes and all probands.

To adjust for family ascertainment, the likelihood of observing the phenotypes and genotypes of each family was computed conditional on observing the phenotypes of the proband, i.e., their cancer status and age of diagnosis or censoring. This ascertainment is justified because all families were ascertained through population-based sampling of probands, and the family history and genotype data could be assumed not to influence the ascertainment.

We computed age-specific breast cancer FRR predicted by the best fitting model using the likelihoods from MENDEL^16^. The FRR for a woman at age *t* with a first-degree relative affected at age *t* was computed as the ratio of two pedigree-likelihoods:

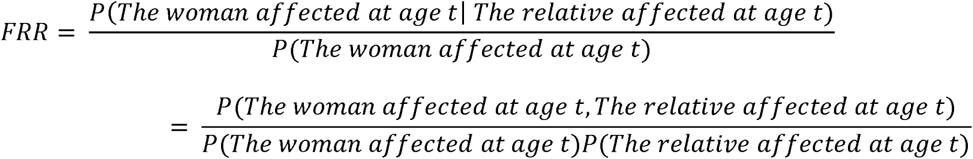

We computed the FRRs separately for women with an affected mother and women with an affected sister.

For *TP53* PV carriers and hypothetical gene PV carriers, we estimated the age-specific cumulative risk (penetrance) to age *t, F(t)*, as

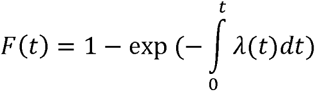

where *λ(t)* is the estimated incidence at age *t* for carriers averaged over the polygenic effects, assuming the UK population incidence for women born in 1940-1949 (the median birth year of the probands was 1948) was used. The 95% confidence interval (CI) of *F(t)* was calculated via a parametric bootstrap: a sample of 10,000 draws was taken from the multivariate normal distribution that the maximum likelihood estimates would be expected to follow under asymptotic likelihood theory; for each age, a corresponding sample of 10,000 cumulative risks was calculated using the formula above, and the 2.5^th^ and 97.5^th^ percentiles of this distribution were taken to be the 95% CI limits.

## RESULTS

A total of 15,022 (86%) probands were screened, and 882 (5.9%) were found to carry a PV in *BRCA1, BRCA2, PALB2, CHEK2, ATM* or *TP53* (Table 1). Of the 1,924 ABCFR relatives tested, 142 from 88 families were found to carry a PV (Table S2). 2,449 (14.1%) and 1,521 (8.7%) probands had a family history of breast cancer in first-degree relatives and second-degree relatives, respectively; the corresponding numbers for a family history of ovarian cancer were 328 (1.9%) and 165 (0.9%), respectively (Table S3).

When a polygenic component only model was fitted, σ_P_^2^(t) was 3.86 (95% CI: 3.27, 4.47) at age 20-29 years (calculated using the middle point, i.e., 25 years; the same for the age ranges below), decreasing to 0.72 (95% CI: 0.42, 1.03) at age 70-79 years (Table S4). After additionally fitting *BRCA1, BRCA2, PALB2, CHEK2* and *ATM*, σ_P_^2^(t) was 2.25 (95% CI: 1.61, 2.88) at age 20-29 years, decreasing to 1.11 (95% CI: 0.70, 1.51) at age 70-79 years. The proportion of the total breast cancer familial variance attributed to these five major genes was therefore 42% at 20-29 years and decreased steadily with age thereafter (Figure 1; Table S5).

**Figure 1.**
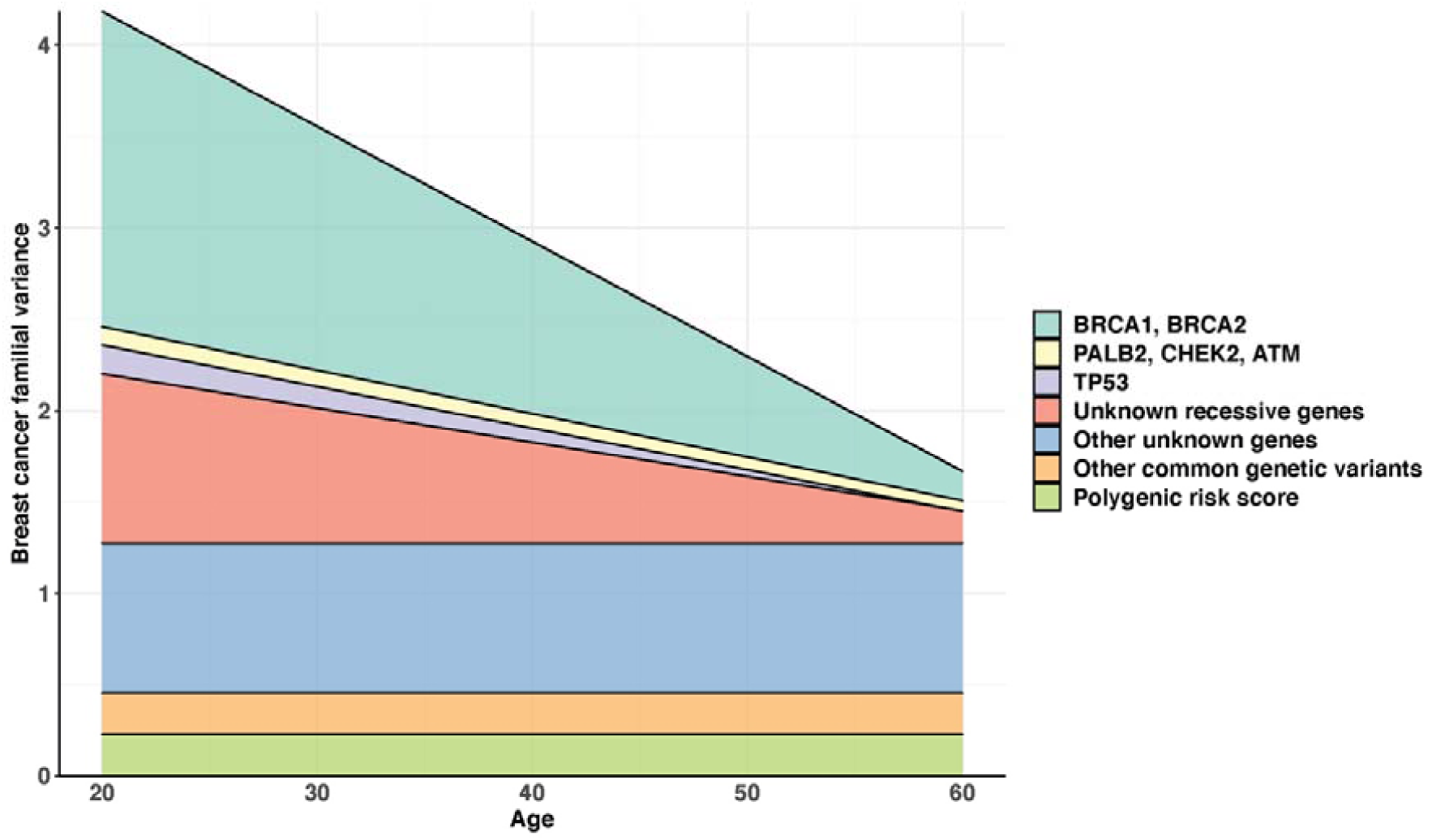
Age-specific breast cancer familial variance explained by genes. For a gene, the age-specific breast cancer familial variance explained by the gene was calculated as the age-specific difference in the σ_P_^2^(t) between the model without the gene and the model with the gene. The variance explained by *BRCA1* and *BRCA2* was the σ_P_^2^(t) of the model including an age-decreasing σ_P_^2^(t) only minus the σ_P_^2^(t) of the model including *BRCA1, BRCA2* and an age-decreasing σ_P_^2^(t). The variance explained by *PALB*2, *CHEK2* and *ATM* was the σ_P_^2^(t) of the model including *BRCA1, BRCA2* and an age-decreasing σ_P_^2^(t) minus the σ_P_^2^(t) of the model including *BRCA1, BRCA2, PALB2, CHEK2, ATM* and an age-decreasing σ_P_^2^(t). The variance explained by *TP53* was the σ_P_^2^(t) of the model including *BRCA1, BRCA2, PALB2, CHEK2, ATM* and an age-decreasing σ_P_^2^(t) minus the σ_P_^2^(t) of the model including *BRCA1, BRCA2, PALB2, CHEK2, ATM, TP53* and an age-decreasing σ_P_^2^(t). The variance explained by unknown recessive genes was the σ_P_^2^(t) of the model including *BRCA1, BRCA2, PALB2, CHEK2, ATM, TP53* and an age-decreasing σ_P_^2^(t) minus the σ_P_^2^(t) of the model including *BRCA1, BRCA2, PALB2, CHEK2, ATM, TP53*, the hypothetical gene and an age-constant σ_P_^2^(t). The variance explained by the polygenic risk score was from Mavaddat et al^11^. The variance explained by other common genetic variants was those explained by all imputable common genetic variants using the OncoArray (i.e., chip heritability), which was approximately twice that explained by known common genetic variants^31^, minus those explained by known common genetic variants, i.e., the polygenic risk score.

We then additionally fitted a sixth hypothetical major gene (Table S4). All three inheritance models (dominant, recessive and general) that included the hypothetical gene gave a better fit than their equivalent nested models without the additional gene (all P<0.03). The dominant inheritance model gave the best fit, and the general inheritance model essentially converged to the dominant inheritance model. Under the dominant inheritance model, the hypothetical gene had a risk allele frequency of 0.003% (95% CI: 0.001%, 0.008%) and an RR of 340 (95% CI: 140, 810); σ _P_^2^(t) was 1.53 (95% CI: 1.37, 1.70), and there was no evidence that σ_P_^2^(t) depended on age (P=0.2).

Most of the difference in log-likelihoods between the dominant inheritance model for the hypothetical gene and its equivalent nested model without the gene was attributed to a small number of families (Figure S1). For all 10 families contributing the most evidence for the hypothetical gene, the proband was diagnosed with breast cancer before age 42 years, and all the affected relatives were diagnosed before age 37 years (Table S6). For three of these families, the proband carried a PV in *TP53*. No PVs in the other genes were identified for the probands of these families from the gene-panel testing data. We therefore hypothesized that the hypothetical gene might reflect the effects of *TP53* PVs, and further extended the models to include *TP53*.

Incorporating *TP53* in addition to the five major genes while fitting an age-decreasing σ _P_^2^(t) improved the model fit (P<10^−15^; Table S7). The best fitting model included a *TP53* RR which decreased linearly with age on the log-RR scale over age 20-49 years and then was constant over age 50-79 years. Under this model, the frequency for *TP53* PVs was 0.017% (95% CI: 0.009%, 0.034%) and the estimated cumulative risk of breast cancer to age 80 years for *TP53* PV carriers was 45.0% (95% CI: 25.5%, 74.0%) (Figure 2). After fitting *TP53*, there was marginal evidence (P=0.07) that σ ^2^(t) decreased with age. Based on the model with an age-decreasing σ_P_^2^(t), the proportion of the total breast cancer familial variance attributed to *TP53* was 3.5% at age 20-29 years and decreased with age thereafter; in terms of the residual familial variance after adjusting for the effects of *BRCA1, BRCA2, PALB2, CHEK2* and *ATM*, the proportion decreased from 6.1% at age 20-29 years (Figure 1; Table S5).

**Figure 2.**
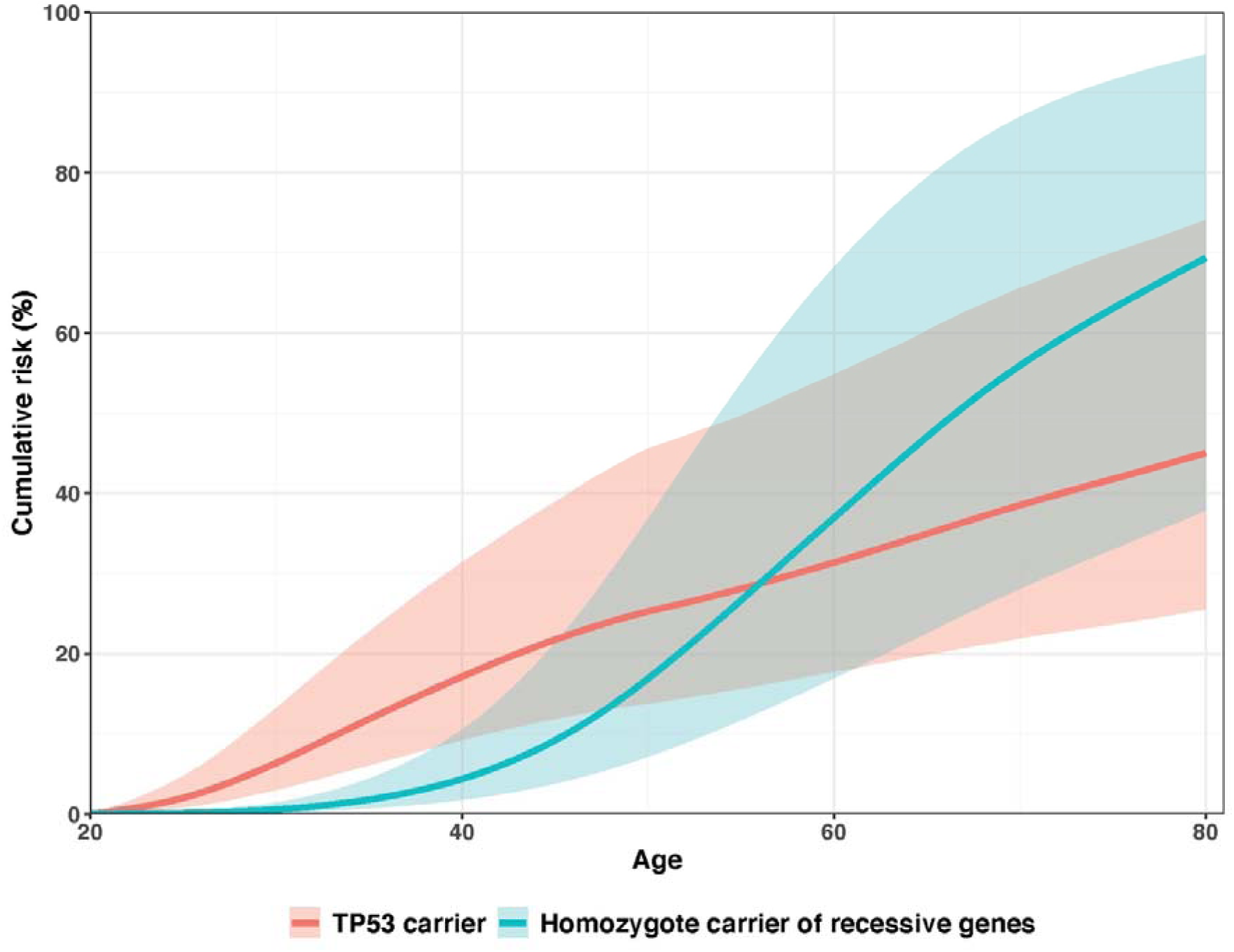
Age-specific cumulative risks with 95% confidence intervals for *TP53* PV carriers and homozygote carriers of PVs in recessive genes. Recessive genes are the hypothetical gene from the best fit model including *BRCA1, BRCA2, PALB2, CHEK2, ATM, TP53*, a seventh hypothetical gene and an age-constant σ_P_^2^(t). Shaded areas are 95% confidence intervals.

Based on the best fitting model that included *TP53* and an age-constant σ_P_^2^(t), we incorporated an additional hypothetical gene (Table 2). There was evidence for the hypothetical gene under both the recessive and general inheritance models (both P<0.02), and the general inheritance model essentially converged to the recessive inheritance model. Under the best fitting recessive inheritance model, the hypothetical gene had a risk allele frequency of 13% (95% CI: 5%, 23%), and an RR of 10 (95% CI: 4, 25) giving a cumulative risk to age 80 of 69.4% (95% CI: 37.9%, 94.7%) for homozygotes (Figure 2). Under this model, σ _P_ ^2^(t) was estimated to be 1.27 (95% CI: 0.94, 1.65) and there was no evidence that it depended on age (P=0.81). The proportion of the total breast cancer familial variance explained by the hypothetical gene was 21.6% at age 20-29 years and decreased steadily with age thereafter; in terms of the residual familial variance after adjusting for the effects of *BRCA1, BRCA2, PALB2, CHEK2, ATM* and *TP53*, the proportion was 39.7% at age 20-29 years and decreased thereafter (Figure 1; Table S5). Under this model, the PV frequencies of *BRCA1, BRCA2, PALB2, CHEK2, ATM* and *TP53* genes were 0.080%, 0.141%, 0.059%, 0.385%, 0.167% and 0.017%, respectively.

**Table 2.**
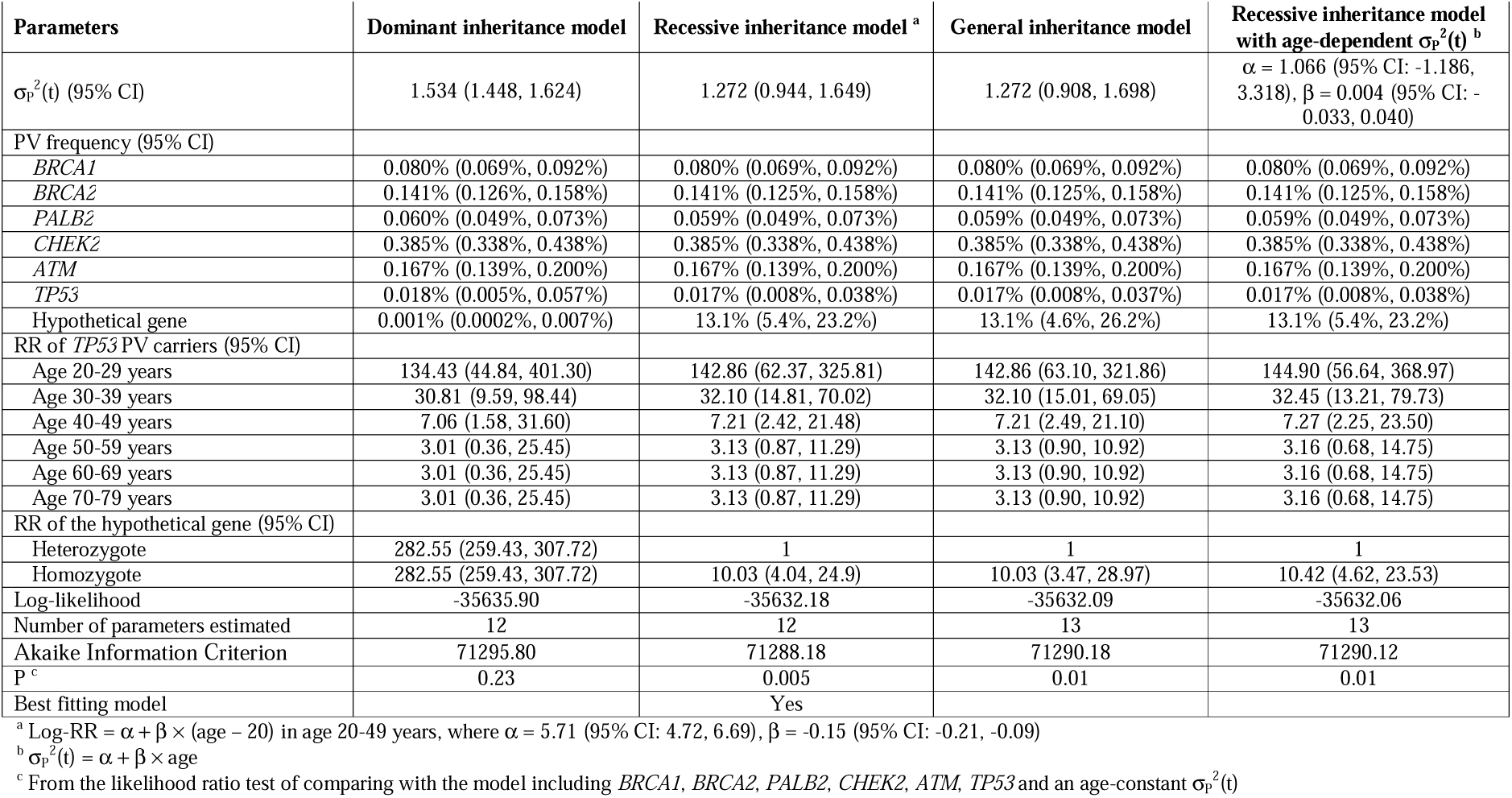
Models including *BRCA1, BRCA2, PALB2, CHEK2, ATM, TP53*, a hypothetical gene and a polygenic component.

| Parameters | Dominant inheritance model | Recessive inheritance model <sup>a</sup> | General inheritance model | Recessive inheritance model with age-dependent $\sigma_P^2(t)$ <sup>b</sup> |
| --- | --- | --- | --- | --- |
| $\sigma_P^2(t)$ (95% CI) | 1.534 (1.448, 1.624) | 1.272 (0.944, 1.649) | 1.272 (0.908, 1.698) | $\alpha = 1.066$ (95% CI: -1.186, 3.318), $\beta = 0.004$ (95% CI: -0.033, 0.040) |
| PV frequency (95% CI) |  |  |  |  |
| *BRCA1* | 0.080% (0.069%, 0.092%) | 0.080% (0.069%, 0.092%) | 0.080% (0.069%, 0.092%) | 0.080% (0.069%, 0.092%) |
| *BRCA2* | 0.141% (0.126%, 0.158%) | 0.141% (0.125%, 0.158%) | 0.141% (0.125%, 0.158%) | 0.141% (0.125%, 0.158%) |
| *PALB2* | 0.060% (0.049%, 0.073%) | 0.059% (0.049%, 0.073%) | 0.059% (0.049%, 0.073%) | 0.059% (0.049%, 0.073%) |
| *CHEK2* | 0.385% (0.338%, 0.438%) | 0.385% (0.338%, 0.438%) | 0.385% (0.338%, 0.438%) | 0.385% (0.338%, 0.438%) |
| *ATM* | 0.167% (0.139%, 0.200%) | 0.167% (0.139%, 0.200%) | 0.167% (0.139%, 0.200%) | 0.167% (0.139%, 0.200%) |
| *TP53* | 0.018% (0.005%, 0.057%) | 0.017% (0.008%, 0.038%) | 0.017% (0.008%, 0.037%) | 0.017% (0.008%, 0.038%) |
| Hypothetical gene | 0.001% (0.0002%, 0.007%) | 13.1% (5.4%, 23.2%) | 13.1% (4.6%, 26.2%) | 13.1% (5.4%, 23.2%) |
| RR of *TP53* PV carriers (95% CI) |  |  |  |  |
| Age 20-29 years | 134.43 (44.84, 401.30) | 142.86 (62.37, 325.81) | 142.86 (63.10, 321.86) | 144.90 (56.64, 368.97) |
| Age 30-39 years | 30.81 (9.59, 98.44) | 32.10 (14.81, 70.02) | 32.10 (15.01, 69.05) | 32.45 (13.21, 79.73) |
| Age 40-49 years | 7.06 (1.58, 31.60) | 7.21 (2.42, 21.48) | 7.21 (2.49, 21.10) | 7.27 (2.25, 23.50) |
| Age 50-59 years | 3.01 (0.36, 25.45) | 3.13 (0.87, 11.29) | 3.13 (0.90, 10.92) | 3.16 (0.68, 14.75) |
| Age 60-69 years | 3.01 (0.36, 25.45) | 3.13 (0.87, 11.29) | 3.13 (0.90, 10.92) | 3.16 (0.68, 14.75) |
| Age 70-79 years | 3.01 (0.36, 25.45) | 3.13 (0.87, 11.29) | 3.13 (0.90, 10.92) | 3.16 (0.68, 14.75) |
| RR of the hypothetical gene (95% CI) |  |  |  |  |
| Heterozygote | 282.55 (259.43, 307.72) | 1 | 1 | 1 |
| Homozygote | 282.55 (259.43, 307.72) | 10.03 (4.04, 24.9) | 10.03 (3.47, 28.97) | 10.42 (4.62, 23.53) |
| Log-likelihood | -35635.90 | -35632.18 | -35632.09 | -35632.06 |
| Number of parameters estimated | 12 | 12 | 13 | 13 |
| Akaike Information Criterion | 71295.80 | 71288.18 | 71290.18 | 71290.12 |
| P <sup>c</sup> | 0.23 | 0.005 | 0.01 | 0.01 |
| Best fitting model |  | Yes |  |  |

<sup>a</sup> Log-RR = $\alpha + \beta \times (\text{age} - 20)$ in age 20-49 years, where $\alpha = 5.71$ (95% CI: 4.72, 6.69), $\beta = -0.15$ (95% CI: -0.21, -0.09)
<sup>b</sup> $\sigma_P^2(t) = \alpha + \beta \times \text{age}$
<sup>c</sup> From the likelihood ratio test of comparing with the model including *BRCA1*, *BRCA2*, *PALB2*, *CHEK2*, *ATM*, *TP53* and an age-constant $\sigma_P^2(t)$

No family contributed a large change in the log-likelihood when the hypothetical recessive gene was added (Figure S1). For all 10 families contributing most to the evidence for the hypothetical gene, the proband had at least one sister diagnosed with breast cancer before or at age 45 years (Table S8). No PV was identified for the probands of these 10 families from the gene-panel testing data.

Similar results were found from the sensitivity analyses in which the PV test sensitivity was reduced to 80% (Table S9). With this sensitivity, the PV frequencies of *BRCA1, BRCA2, PALB2, CHEK2, ATM* and *TP53* genes were 0.089%, 0.157%, 0.067%, 0.432%, 0.187% and 0.020%, respectively.

The age-specific FRRs associated with a family history in a first-degree relative predicted by the best fitting recessive inheritance model were consistent with those observed by the largest combined analysis of epidemiological studies^1^ (Table 3). At each age, the FRR associated with a sister affected was slightly greater than the FRR associated with the mother affected.

**Table 3.**
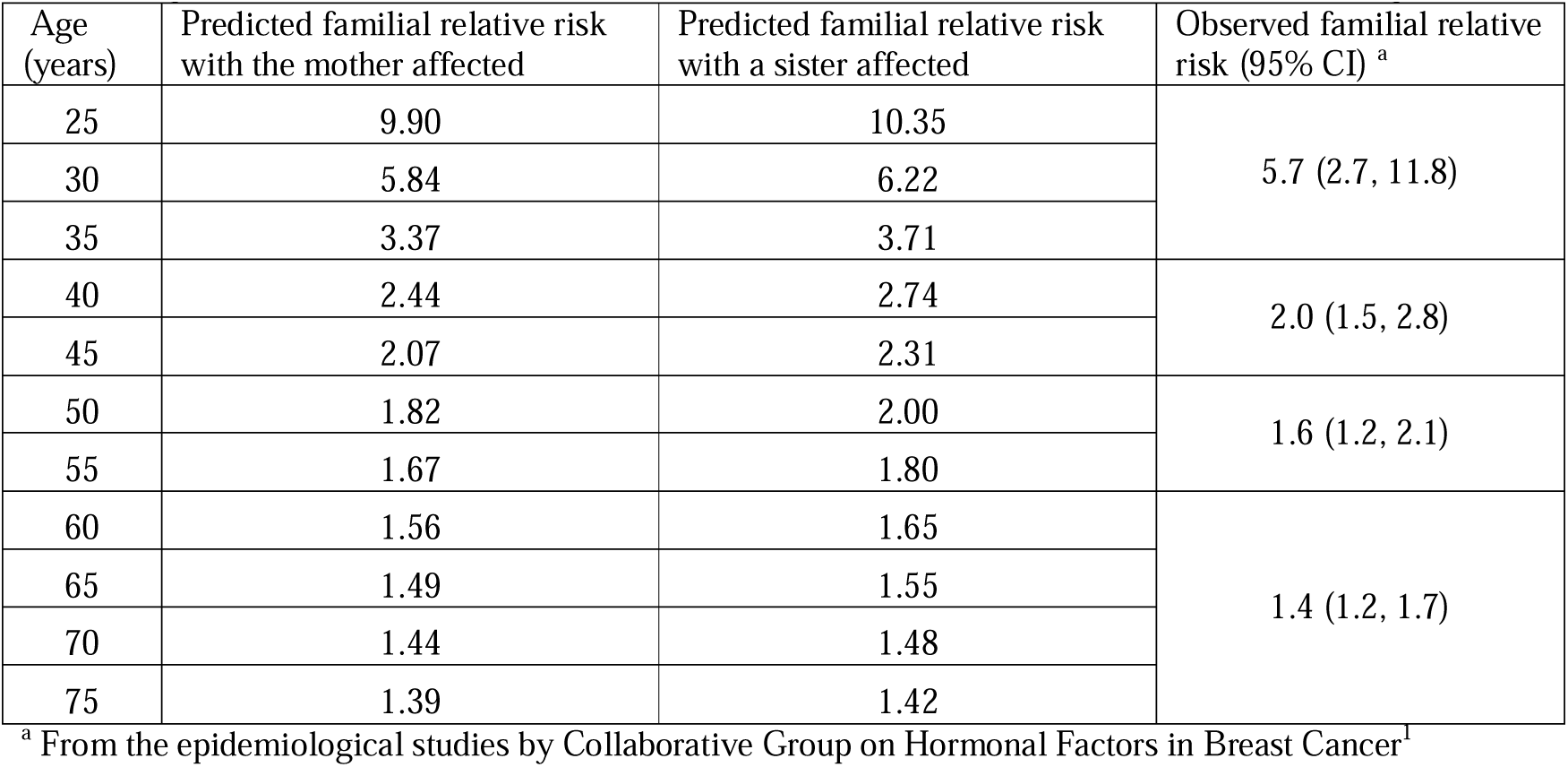
Age-specific breast cancer familial relative risks associated with an affected first-degree relative.

## DISCUSSION

This study provides new insights into the genetic susceptibility of breast cancer. In terms of explaining why women of the same age differ in risk, the known susceptibility genes including *BRCA1, BRCA2, PALB2, CHEK2, ATM* and *TP53* play more important roles at younger ages; the proportion of the total breast cancer familial variance explained by these genes was 46% at age 20-29 years and decreased steadily with age thereafter. GWAS have identified several hundred common genetic variants associated with breast cancer risk^31^; these variants combine multiplicatively, and their effects can thus be summarised by a PRS. The most extensively validated PRS, comprising 313 common genetic variants, is associated with breast cancer risk with little evidence of variation in the relative risk by age^11^, implying that the PRS explains a higher fraction of the breast cancer familial variance at older ages than at younger ages^32,33^. The odds ratio (OR) per standard deviation of the PRS is 1.61, equivalent to a familial variance of ∼0.23. Therefore, the PRS accounts for about 20% of the residual polygenic variance of 1.27 estimated after taking into account the known and predicted additional major gene(s), in line with previous estimates for the PRS contribution^11,20^. The breast cancer familial variance explained by all imputable common genetic variants using the OncoArray (i.e., chip heritability) was estimated to be approximately twice that explained by the known common genetic variants (and thus about 40% of the residual polygenic variance)^31^. As illustrated in Figure 1, there is substantial breast cancer familial variance that cannot be explained by the known susceptibility genes and common genetic variants, and the unexplained familial variance is larger at younger ages. Therefore, breast cancer genomic studies focused on cases diagnosed at younger ages may be a fruitful approach to identifying novel breast cancer genetic susceptibility genes or variants. Our analyses predicted a recessively inherited risk component explaining a substantial proportion of the residual familial variance after taking into account PVs in *BRCA1, BRCA2, PALB2, CHEK2, ATM, TP53* and the PRS, with the proportion greater at younger ages (44% at ages 20-29 years) and decreased steadily with age thereafter.

Although our study sample overlapped with the data used in three previous analyses^12-14^, the current dataset was seven times larger. Moreover, we were able to include PV data for the large majority of probands, providing much more robust analyses. Our evidence for an additional recessive major breast cancer susceptibility gene, after considering the effects of PVs in *BRCA1, BRCA2, PALB2, CHEK2, ATM* and *TP53*, is consistent with the results from previous analyses^12-15^. The best fitting model predicted age-specific FRRs consistent with the observed FRRs by epidemiological studies. The recessive risk component leads to the prediction that the FRR will be higher for sisters of affected women than for mothers of affected daughters, and this is consistent with observations in population-based epidemiological studies^1,13,25,34-36^. Although the lower relative risk for mothers could be partly explained by them being parous by definition, the risk difference is more profound before age 50 years when mothers are pre-menopausal and the protective effect of parity is weaker^1,35^. Our analysis used birth-cohort-specific incidences; therefore, the results are unlikely to be due to the higher incidence for sisters, who were born more recently than mothers. However, we cannot rule out the possibility that part of the increased risk to sisters may be due to surveillance, since sisters of breast cancer cases are more likely to participate in screening programs.

We also cannot rule out the role of non-genetic familial factors in explaining familial aggregation of breast cancer as we only considered genetic models. The Nordic Twin Study of breast cancer found evidence for non-genetic effects shared by twins, especially at younger ages, which would also be consistent with the FRR being greater for sister pairs than mother-daughter pairs^30^ and could reflect factors operating prior to adulthood such as puberty-related risk factors.

Our results also do not necessarily imply there is a single additional susceptibility gene – indeed this is highly unlikely since such a gene would probably have been identified through linkage or association studies. It is more likely that the recessive component reflects the combined effects of multiple variants (and in more than one gene or non-coding region), each potentially being associated with different effects. Nevertheless, our results could inform the design of sequencing studies to try to identify such variants by focussing on the analysis of families with multiple affected siblings, and which do not have PVs in the known genes.

Breast cancer FRR depends on pathological subtype, and the risks associated with both known breast cancer susceptibility genes and PRS differ across subtypes^11,34,37^. Further analyses of datasets with breast cancer subtype data are needed to investigate the extent to which the recessive risk component and the residual polygenic variance are subtype-specific.

We calculated the age-specific proportion of breast cancer familial variance explained by PVs in major genes as the difference in the polygenic variance between nested models. The results in Table S5 suggest the proportion at age 60 and older might be minimal, despite the fact the RRs for breast cancer associated with PVs at those ages are greater than 1. The estimated contribution of PVs at older ages may have been underestimated due to the fact that the total breast cancer familial variance used to calculate the contributions was based on a model without any major genes. Under this model, the polygenic variance decreased linearly with age, which may have resulted in imprecision of the estimated variance at older ages.

We previously conducted a population-based study of women with breast cancer diagnosed at a young age in ABCFR and confirmed that germline *TP53* PVs occur among women diagnosed at a very age^38^. Here, we estimated the PV frequency to be 0.017%, similar to the 1/5000 estimated by a segregation analysis of 278 breast cancer families ascertained via population-based cases diagnosed before age 30 years^39^. Reliable breast cancer risk estimates associated with *TP53* PVs are lacking. Two studies of sarcoma families reported a breast cancer cumulative risk for *TP53* PV carriers of approximately 37% to age 80 years and 54% to age 70 years, respectively^40,41^; however, these studies did not report confidence intervals. Our study estimated age-specific breast cancer risk and confidence intervals for *TP53* PV carriers; however, the number of carriers was small, and the confidence intervals were correspondingly wide.

The findings of the current study can be used to update breast cancer risk models, in particular BOADICEA which currently considers the effects of PVs in known major genes and a polygenic component^21^. Thus, the model could be updated by incorporating the recessive component, using the revised estimate for the polygenic variance, and adding *TP53* PVs. However, the extent to which including the additional genes and using the updated polygenic variance improves breast cancer risk prediction overall needs to be investigated. Moreover, such a revised model would need to be validated using independent studies.

To allow for the possibility that not all PVs can be detected by the test methods used, our analysis included a test sensitivity parameter, which was assumed to be ∼90%. The actual sensitivity is difficult to estimate and will depend on the methods used. The current implementation of BOADICEA assumes sensitivities of 89%, 96%, 92%, 98% and 94% for *BRCA1, BRCA2, PALB2, CHEK2* and *ATM*, respectively, based on typical clinically testing approaches^2,21^, but the sensitivity will be lower for research testing, particularly since large rearrangements would not have been detected in the targeted sequencing used in SEARCH^2^. Using lower test sensitivities might have resulted in some underestimation of the polygenic component and some overestimation of the contributions of rare PVs to familial aggregation, through the estimation of higher PV frequencies. However, the differences would be small, as our estimates of PV frequencies for *BRCA1* and *BRCA2* were slightly greater than, and the estimates for *PALB2, CHEK2* and *ATM* were similar to, those assumed in the BOADICEA which had been derived from large population-based targeted-sequencing data and adjusted for the test sensitivity of targeted-sequencing^21^. On the other hand, by using lower test sensitivities, our analysis reduced the possibility that the recessive risk component is simply due to unidentified variants in any of the considered major genes; our more conservative sensitivity analysis assuming the test sensitivity to be 80% still provided evidence for a recessive risk component.

Our study has several strengths. First, it included data from more than 17,000 three-generational families ascertained via probands from population-based studies and screened for high- and moderate-risk PVs in the major known susceptibility genes. This is by far the largest of its kind to date. Second, our study is the first to incorporate the explicit age-specific effects of PVs in *PALB2, CHEK2, ATM* and *TP53* in addition to *BRCA1* and *BRCA2*, while modelling the residual familial variance of breast cancer as a function of age using a polygenic component.

The study has also some limitations. First, cancer family history in some families was self-reported and therefore subject to reporting errors, though reporting of breast cancer in first-degree relatives is generally considered to be accurate^42^. Second, we modelled the major genes using a single locus with eight alleles and assumed the genes were in a dominant hierarchy, rather than seven loci each with two alleles. While this approach substantially reduces the computational time, it could introduce some imprecision in the parameter estimates, although the impact is likely to be minimal because the PVs in the known genes are rare. The polygenic component was also approximated using a binomial distribution inherited under the hypergeometric model; previous analyses had found that results are insensitive to the number of loci assumed^12^, but this might not be true for more complex models.

In conclusion, by considering the explicit effects of established major breast cancer susceptibility genes and polygenes and using the largest sample size of its kind, our analysis estimates the proportion of breast cancer familial aggregation that explained by established susceptibility genes and variants, and provides evidence for an additional recessive risk component which could explain a substantial proportion of the residual familial aggregation, especially at a younger age. Our findings are informative for the design of sequencing studies to identify novel breast cancer susceptibility genes, and modelling breast cancer genetic susceptibility for disease risk prediction.

## Data Availability

Requests for access to the data analysed in this study should be made to J.L.H. (ABCFR) and P.D.P. (SEARCH).

## ACKNOWLEDGMENTS

We thank all the participants in this study, the entire team of the ABCFR and past and current investigators. The work was supported by Cancer Research UK grant PPRPGM-Nov20\100002, NIHR Cambridge Biomedical Research Centre, Victorian Cancer Agency grant ECRF19020, Picchi Award for Excellence in Cancer Research from the Victorian Comprehensive Cancer Centre and the U.S. National Institute of Health (grant number RO1CA159868). The ABCFR was supported in Australia by the National Health and Medical Research Council (NHMRC), the New South Wales Cancer Council, the Victorian Health Promotion Foundation, the Victorian Breast Cancer Research Consortium, Cancer Australia, and the National Breast Cancer Foundation. The six sites of the Breast Cancer Family Registry (BCFR) were supported by grant UM1 CA164920 from the U.S. National Cancer Institute. The content of this manuscript does not necessarily reflect the views or policies of the National Cancer Institute or any of the collaborating centres in the BCFR, nor does mention of trade names, commercial products, or organizations imply endorsement by the U.S. Government or the BCFR. Sequencing of the ABCFR samples was supported by an NHMRC Program grant (APP1074383), The National Breast Cancer Foundation (BRA-STRAP; NT-15-016), NHMRC European Union Collaborative Research Grant (APP1101400) and Monash University, Melbourne, Australia. SEARCH was funded by Cancer Research UK (C490/A16561) and the NIHR Biomedical Research Centre at the University of Cambridge and the University of Cambridge has received salary support for P.D.P.P. from the NHS in the East of England through the Clinical Academic Reserve. Sequencing of the SEARCH samples was funded by European Union’s Horizon 2020 Research and Innovation Programme (BRIDGES: grant number 634935). S.L. is a Victorian Cancer Agency (VCA) Early Career Research Fellow (ECRF19020). TN-D is a VCA Mid-Career Research Fellow (MCRF21029). M.C.S. is an NHMRC Senior Research Fellow (APP1155163). J.L.H. is an NHMRC Senior Principal Research Fellow.

This work was performed using resources provided by the Cambridge Service for Data Driven Discovery (CSD3) operated by the University of Cambridge Research Computing Service (www.csd3.cam.ac.uk), provided by Dell EMC and Intel using Tier-2 funding from the Engineering and Physical Sciences Research Council (capital grant EP/T022159/1), and DiRAC funding from the Science and Technology Facilities Council (www.dirac.ac.uk).

## DATA AND CODE AVAILABILITY

Requests for access to the data analysed in this study should be made to J.L.H. (ABCFR) and P.D.P. (SEARCH). Requests for access to the analytical code should be made to S.L. and A.C.A..

