## Supplementary tables and figures for "Segregation analysis of 17,425 population-based breast cancer families: evidence for genetic susceptibility and risk prediction"

### Table S1 Assumed sensitivity of the pathogenic variant test methods in the ABCFR

| Method | Assumed sensitivity |
| --- | --- |
| Heteroduplex Analysis | 70% |
| Protein Truncation Test (PTT) | 90% |
| Predictive Sequencing | 100% |
| 2-Dimensional Gel Electrophoresis | 70% |
| Automated DNA Sequencing | 100% |
| Multiplex ligation-dependent probe amplification (MLPA) | 100% |
| MYRIAD DNA Sequencing | 100% |
| Manual DNA Sequencing | 100% |
| Denaturing High Performance Liquid (DHPLC) | 90% |
| PCR and Agarose Gel Electrophoresis | 70% |
| 5’Nuclease Assay (TaqMan) | 95% |
| Gene-panel test | 100% |

The test sensitivity used in analysis was the sum of the sensitivities of the test methods used, weighted by the proportional lengths of the exons screened, multiplying by 90% if MLPA had not been conducted. If MLPA had been conducted, 10% multiplying the sensitivity of MLPA weighted by the proportional lengths of the exons screened was added.

### Table S2 Number of ABCFR relatives found to be carriers by the diagnosis age and PV status of the proband a

| **Gene** | **Number of relatives and families** | | **Age groups of the probands (years)** | | | | | | | | | | | |
| --- | --- | --- | --- | --- | --- | --- | --- | --- | --- | --- | --- | --- | --- | --- |
|  |  |  | **Case families** | | | | | | **Control families** | | | | | |
|  |  |  | **<30** | **30-39** | **40-49** | **50-59** | **60-69** | **Total** | **<30** | **30-39** | **40-49** | **50-59** | **60-69** | **Total** |
| *BRCA1* | Number of relatives | | 1 | 27 | 22 | 3 | 0 | 53 | 0 | 1 | 2 | 0 | 0 | 3 |
|  | Number of families where the relatives come from | Probands carriers | 1 | 17 | 9 | 1 | 0 | 28 | 0 | 1 | 1 | 0 | 0 | 2 |
|  |  | Probands non-carriers | 0 | 0 | 1 | 1 | 0 | 2 | 0 | 0 | 0 | 0 | 0 | 0 |
|  |  | Probands untested | 0 | 0 | 0 | 0 | 0 | 0 | 0 | 0 | 0 | 0 | 0 | 0 |
| *BRCA2* | Number of relatives | | 4 | 25 | 7 | 5 | 0 | 41 | 0 | 0 | 0 | 0 | 0 | 0 |
|  | Number of families where the relatives come from | Probands carriers | 3 | 14 | 2 | 0 | 0 | 19 | 0 | 0 | 0 | 0 | 0 | 0 |
|  |  | Probands non-carriers | 0 | 0 | 3 | 2 | 0 | 5 | 0 | 0 | 0 | 0 | 0 | 0 |
|  |  | Probands untested | 0 | 2 | 0 | 0 | 0 | 2 | 0 | 0 | 0 | 0 | 0 | 0 |
| *PALB2* | Number of relatives | | 3 | 2 | 7 | 1 | 0 | 13 | 0 | 0 | 0 | 0 | 0 | 0 |
|  | Number of families where the relatives come from | Probands carriers | 1 | 1 | 3 | 1 | 0 | 6 | 0 | 0 | 0 | 0 | 0 | 0 |
|  |  | Probands non-carriers | 0 | 0 | 2 | 0 | 0 | 2 | 0 | 0 | 0 | 0 | 0 | 0 |
|  |  | Probands untested | 0 | 0 | 0 | 0 | 0 | 0 | 0 | 0 | 0 | 0 | 0 | 0 |
| *CHEK2* | Number of relatives | | 4 | 6 | 6 | 2 | 0 | 18 | 0 | 0 | 0 | 0 | 0 | 0 |
|  | Number of families where the relatives come from | Probands carriers | 2 | 4 | 4 | 0 | 0 | 10 | 0 | 0 | 0 | 0 | 0 | 0 |
|  |  | Probands non-carriers | 0 | 1 | 0 | 2 | 0 | 3 | 0 | 0 | 0 | 0 | 0 | 0 |
|  |  | Probands untested | 0 | 0 | 0 | 0 | 0 | 0 | 0 | 0 | 0 | 0 | 0 | 0 |
| *ATM* | Number of relatives | | 0 | 3 | 3 | 7 | 1 | 14 | 0 | 1 | 0 | 1 | 0 | 2 |
|  | Number of families where the relatives come from | Probands carriers | 0 | 0 | 1 | 2 | 1 | 4 | 0 | 0 | 0 | 1 | 0 | 1 |
|  |  | Probands non-carriers | 0 | 3 | 2 | 2 | 0 | 7 | 0 | 1 | 0 | 0 | 0 | 1 |
|  |  | Probands untested | 0 | 0 | 0 | 0 | 0 | 0 | 0 | 0 | 0 | 0 | 0 | 0 |

^a^ 2 relatives had a PV in two genes: case families – one relative (proband aged 30-39 years) in *BRCA2* and *CHEK2*; control families – one relative (proband aged 30-39 years) in *BRCA1* and *ATM*

### Table S3 Number of relatives diagnosed with breast or ovarian cancer by study and the diagnosis age and PV status of the proband

| **PV status** | **Relative type** | **Cancer site** | **ABCFR case families** | | | | | |  | **ABCFR control families** | | | | | |  | **SEARCH** | | | | | | |
| --- | --- | --- | --- | --- | --- | --- | --- | --- | --- | --- | --- | --- | --- | --- | --- | --- | --- | --- | --- | --- | --- | --- | --- |
|  |  |  | **Age group (years)** | | | | | **Total** |  | **Age group (years)** | | | | | **Total** |  | **Age group (years)** | | | | | | **Total** |
|  |  |  | **<30** | **30-39** | **40-49** | **50-59** | **60-69** |  |  | **<30** | **30-39** | **40-49** | **50-59** | **60-69** |  |  | **<30** | **30-39** | **40-49** | **50-59** | **60-69** | **70-79** |  |
| *BRCA1* PV carriers | 1st-degree | Breast | 0 | 27 | 7 | 3 | 0 | 37 |  | 0 | 0 | 2 | 0 | 0 | 2 |  | 2 | 6 | 14 | 12 | 4 | 0 | 38 |
|  |  | Ovary | 0 | 4 | 3 | 0 | 0 | 7 |  | 0 | 0 | 0 | 0 | 0 | 0 |  | 0 | 3 | 4 | 5 | 4 | 0 | 16 |
|  | 2^nd^-degree | Breast | 4 | 17 | 5 | 0 | 0 | 26 |  | 0 | 1 | 1 | 0 | 0 | 2 |  | 1 | 5 | 9 | 3 | 1 | 0 | 19 |
|  |  | Ovary | 0 | 6 | 1 | 0 | 0 | 7 |  | 0 | 0 | 1 | 0 | 0 | 1 |  | 0 | 2 | 5 | 1 | 0 | 0 | 8 |
| *BRCA2* PV carriers | 1st-degree | Breast | 1 | 9 | 1 | 1 | 0 | 12 |  | 0 | 1 | 0 | 0 | 0 | 1 |  | 0 | 7 | 25 | 34 | 15 | 0 | 81 |
|  |  | Ovary | 0 | 0 | 0 | 1 | 0 | 1 |  | 0 | 1 | 0 | 0 | 0 | 1 |  | 0 | 1 | 4 | 8 | 3 | 0 | 16 |
|  | 2^nd^-degree | Breast | 6 | 11 | 3 | 1 | 0 | 21 |  | 0 | 1 | 0 | 0 | 0 | 1 |  | 1 | 11 | 9 | 7 | 1 | 0 | 29 |
|  |  | Ovary | 0 | 1 | 0 | 0 | 0 | 1 |  | 1 | 1 | 0 | 0 | 0 | 2 |  | 0 | 2 | 1 | 1 | 1 | 0 | 5 |
| *PALB2* PV carriers | 1st-degree | Breast | 0 | 3 | 1 | 2 | 0 | 6 |  | 0 | 0 | 0 | 0 | 0 | 0 |  | 0 | 0 | 3 | 8 | 6 | 0 | 17 |
|  |  | Ovary | 0 | 0 | 0 | 0 | 0 | 0 |  | 0 | 0 | 0 | 0 | 0 | 0 |  | 0 | 0 | 1 | 1 | 0 | 0 | 2 |
|  | 2^nd^-degree | Breast | 3 | 2 | 1 | 2 | 0 | 8 |  | 0 | 1 | 0 | 0 | 0 | 1 |  | 0 | 0 | 3 | 0 | 1 | 0 | 4 |
|  |  | Ovary | 0 | 0 | 0 | 1 | 0 | 1 |  | 0 | 0 | 0 | 0 | 0 | 0 |  | 0 | 0 | 1 | 0 | 0 | 0 | 1 |
| *CHEK2* PV carriers | 1st-degree | Breast | 0 | 1 | 0 | 1 | 0 | 2 |  | 0 | 0 | 2 | 0 | 0 | 2 |  | 1 | 0 | 9 | 22 | 8 | 0 | 40 |
|  |  | Ovary | 0 | 0 | 0 | 0 | 0 | 0 |  | 0 | 0 | 0 | 0 | 0 | 0 |  | 0 | 0 | 0 | 2 | 2 | 0 | 4 |
|  | 2^nd^-degree | Breast | 2 | 4 | 0 | 0 | 0 | 6 |  | 0 | 0 | 0 | 0 | 0 | 0 |  | 1 | 1 | 5 | 6 | 3 | 0 | 16 |
|  |  | Ovary | 1 | 0 | 1 | 0 | 0 | 2 |  | 0 | 0 | 0 | 0 | 0 | 0 |  | 0 | 0 | 1 | 1 | 1 | 0 | 3 |
| *ATM* PV carriers | 1st-degree | Breast | 0 | 0 | 0 | 0 | 0 | 0 |  | 0 | 0 | 0 | 0 | 0 | 0 |  | 0 | 0 | 7 | 13 | 7 | 0 | 27 |
|  |  | Ovary | 0 | 0 | 0 | 0 | 0 | 0 |  | 0 | 0 | 0 | 0 | 0 | 0 |  | 0 | 0 | 1 | 2 | 3 | 0 | 6 |
|  | 2^nd^-degree | Breast | 0 | 1 | 2 | 0 | 0 | 3 |  | 0 | 1 | 0 | 0 | 0 | 1 |  | 0 | 0 | 6 | 0 | 3 | 0 | 9 |
|  |  | Ovary | 0 | 0 | 0 | 0 | 0 | 0 |  | 0 | 0 | 0 | 0 | 0 | 0 |  | 0 | 0 | 0 | 0 | 1 | 0 | 1 |
| *TP53* PV carriers | 1st-degree | Breast | 0 | 2 | 0 | 1 | 0 | 3 |  | 0 | 0 | 0 | 0 | 0 | 0 |  | 1 | 0 | 3 | 0 | 0 | 0 | 4 |
|  |  | Ovary | 1 | 0 | 0 | 0 | 0 | 1 |  | 0 | 0 | 0 | 0 | 0 | 0 |  | 0 | 0 | 0 | 0 | 0 | 0 | 0 |
|  | 2^nd^-degree | Breast | 0 | 9 | 0 | 1 | 0 | 10 |  | 0 | 0 | 0 | 0 | 0 | 0 |  | 0 | 0 | 1 | 0 | 0 | 0 | 1 |
|  |  | Ovary | 0 | 0 | 0 | 0 | 0 | 0 |  | 0 | 0 | 0 | 0 | 0 | 0 |  | 0 | 0 | 0 | 0 | 0 | 0 | 0 |
| Non-PV carriers | 1st-degree | Breast | 8 | 63 | 39 | 45 | 4 | 159 |  | 2 | 16 | 17 | 14 | 8 | 57 |  | 3 | 64 | 353 | 717 | 676 | 1 | 1814 |
|  |  | Ovary | 0 | 10 | 6 | 5 | 1 | 22 |  | 0 | 7 | 6 | 4 | 0 | 17 |  | 1 | 7 | 44 | 87 | 62 | 0 | 201 |
|  | 2^nd^-degree | Breast | 15 | 184 | 97 | 94 | 12 | 402 |  | 10 | 92 | 48 | 36 | 9 | 195 |  | 1 | 65 | 217 | 301 | 169 | 1 | 754 |
|  |  | Ovary | 1 | 16 | 6 | 8 | 0 | 31 |  | 3 | 7 | 5 | 2 | 1 | 18 |  | 0 | 8 | 23 | 28 | 11 | 0 | 70 |
| Untested | 1st-degree | Breast | 1 | 11 | 3 | 0 | 2 | 17 |  | 0 | 3 | 1 | 0 | 1 | 5 |  | 1 | 25 | 91 | 137 | 83 | 0 | 337 |
|  |  | Ovary | 0 | 1 | 0 | 0 | 0 | 1 |  | 0 | 0 | 0 | 0 | 0 | 0 |  | 1 | 4 | 12 | 13 | 7 | 0 | 37 |
|  | 2^nd^-degree | Breast | 2 | 21 | 2 | 4 | 0 | 29 |  | 2 | 7 | 3 | 4 | 5 | 21 |  | 2 | 20 | 33 | 45 | 22 | 0 | 122 |
|  |  | Ovary | 1 | 2 | 0 | 0 | 0 | 3 |  | 0 | 2 | 0 | 0 | 1 | 3 |  | 0 | 0 | 7 | 1 | 2 | 0 | 10 |

### Table S4 Models including BRCA1, BRCA2, PALB2, CHEK2, ATM, a sixth hypothetical gene and a polygenic component

| **Parameter** | **Polygenic component only** | ***BRCA1, BRCA2* and a polygenic component** | ***BRCA1, BRCA2, PALB2, CHEK2, ATM* and a polygenic component** | **With a sixth hypothetical gene** | | | |
| --- | --- | --- | --- | --- | --- | --- | --- |
|  |  |  |  | **Dominant inheritance model** | **Recessive inheritance model** | **General inheritance model** | **Dominant inheritance model with an age-constant σ_P_^2^(t)** |
| σ_P_^2^(t) (95% CI) ^a^ | α = 5.442 (95% CI: 4.447, 6.438), β = -0.062 (95% CI: -0.079, -0.046) | α = 2.934 (95% CI: 1.854, 4.015), β = -0.024 (95% CI: -0.043, -0.005) | α = 2.814 (95% CI: 1.698, 3.929), β = -0.023 (95% CI: -0.042, -0.003) | α = 2.303 (95% CI: 1.133, 3.472), β = -0.014 (95% CI: -0.035, 0.007) | α = 1.397 (95% CI: 0.257, 2.537), β = -0.002 (95% CI: -0.022, 0.018) | α = 2.291 (95% CI: 1.121, 3.460), β = -0.014 (95% CI: -0.035, 0.007) | 1.530  (1.373, 1.696) |
| PV frequency (95% CI) |  |  |  |  |  |  |  |
| *BRCA1* | N/A | 0.082%  (0.071%, 0.094%) | 0.081%  (0.070%, 0.093%) | 0.080%  (0.069%, 0.092%) | 0.080%  (0.069%, 0.093%) | 0.080%  (0.069%, 0.092%) | 0.080%  (0.069%, 0.092%) |
| *BRCA2* | N/A | 0.142%  (0.126%, 0.159%) | 0.141%  (0.126%, 0.159%) | 0.141%  (0.126%, 0.159%) | 0.141%  (0.126%, 0.158%) | 0.141%  (0.126%, 0.159%) | 0.141%  (0.126%, 0.159%) |
| *PALB2* | N/A | N/A | 0.060%  (0.049%, 0.073%) | 0.060%  (0.049%, 0.073%) | 0.060%  (0.049%, 0.073%) | 0.060%  (0.049%, 0.073%) | 0.060%  (0.049%, 0.073%) |
| *CHEK2* | N/A | N/A | 0.385%  (0.338%, 0.438%) | 0.385%  (0.338%, 0.439%) | 0.385%  (0.338%, 0.438%) | 0.385%  (0.338%, 0.439%) | 0.385%  (0.338%, 0.438%) |
| *ATM* | N/A | N/A | 0.167%  (0.139%, 0.200%) | 0.167%  (0.139%, 0.200%) | 0.167%  (0.139%, 0.200%) | 0.167%  (0.139%, 0.200%) | 0.167%  (0.139%, 0.200%) |
| Hypothetical gene | N/A | N/A | N/A | 0.002%  (0.001%, 0.006%) | 10.6%  (0.5%, 21.1%) | 0.003%  (0.001%, 0.006%) | 0.003%  (0.001%, 0.008%) |
| RR of hypothetical gene (95% CI) |  |  |  |  |  |  |  |
| Heterozygote | N/A | N/A | N/A | 415.87  (283.55, 609.93) | 1 | 398.63  (168.71, 941.89) | 340.15  (143.03, 808.94) |
| Homozygote | N/A | N/A | N/A | 415.87  (283.55, 609.93) | 11.61  (4.25, 31.71) | 36641.33  (0, 1.3 × 10^16^) | 340.15  (143.03, 808.94) |
| Log-likelihood | -48094.17 | -46507.53 | -35414.41 | -35406.04 | -35410.62 | -35406.05 | -35406.76 |
| Number of parameters estimated | 2 | 4 | 7 | 9 | 9 | 10 | 8 |
| Akaike Information Criterion | 96192.34 | 93031.06 | 70842.82 | 70830.08 | 70839.24 | 70832.10 | 70829.52 |
| P ^b^ | N/A | N/A | N/A | 2.3 × 10^-4^ | 0.02 | 8.1 × 10^-4^ | 9.2 × 10^-5^ |
| Best fitting model |  |  |  |  |  |  | Yes |

^a^ σ_P_^2^(t) = α + β × age

^b^ From the likelihood ratio test of comparing with the model including *BRCA1*, *BRCA2*, *PALB2*, *CHEK2*, *ATM* and an age-dependent σ_P_^2^(t)

### Table S5 Age-specific polygenic variance after fitting major genes and age-specific proportion of breast cancer familial variance explained by major genes

| **Age group (years)** | **σ_P_^2^(t) (95% CI) ^a^** | | | | | **Proportion of breast cancer familial variance explained by major genes ^d^** | | | |
| --- | --- | --- | --- | --- | --- | --- | --- | --- | --- |
|  | **Only a polygenic component fitted ^b^** | ***BRCA1, BRCA2* fitted ^b^** | ***BRCA1, BRCA2, PALB2, CHEK2, ATM* fitted ^b^** | ***BRCA1, BRCA2, PALB2, CHEK2, ATM, TP53* fitted ^b^** | ***BRCA1, BRCA2, PALB2, CHEK2, ATM, TP53*, a seventh hypothetical gene fitted ^c^** | ***BRCA1, BRCA2*** | ***BRCA1, BRCA2,***  ***PALB2, CHEK2, ATM*** | ***TP53*** | **The seventh hypothetical gene** |
| 20-29 | 3.869  (3.274, 4.465) | 2.340  (1.723, 2.959) | 2.245  (1.609, 2.883) | 2.108  (1.472, 2.745) | 1.272  (0.944, 1.649) | 39.52% | 41.98% | 3.54% | 21.59% |
| 30-39 | 3.240  (2.805, 3.679) | 2.102  (1.670, 2.538) | 2.017  (1.572, 2.466) | 1.920  (1.474, 2.370) | 1.272  (0.944, 1.649) | 35.12% | 37.74% | 2.99% | 20.00% |
| 40-49 | 2.611  (2.611, 2.902) | 1.864  (1.599, 2.131) | 1.790  (1.518, 2.063) | 1.733  (1.460, 2.008) | 1.272  (0.944, 1.649) | 28.60% | 31.46% | 2.17% | 17.65% |
| 50-59 | 1.982  (1.803, 2.162) | 1.626  (1.462, 1.789) | 1.562  (1.398, 1.724) | 1.546  (1.380, 1.709) | 1.272  (0.944, 1.649) | 17.95% | 21.19% | 0.83% | 13.79% |
| 60-69 | 1.353  (1.164, 1.543) | 1.389  (1.152, 1.626) | 1.334  (1.096, 1.573) | 1.358  (1.121, 1.596) | 1.272  (0.944, 1.649) | 0% | 1.37% | 0% | 6.35% |
| 70-79 | 0.724  (0.417, 1.032) | 1.151  (0.753, 1.550) | 1.107  (0.701, 1.514) | 1.171  (0.768, 1.575) | 1.272  (0.944, 1.649) | 0% | 0% | 0% | 0% |

^a^ For each age group, the variance was assumed to be the variance at the middle point age

^b^ From the model in which σ_P_^2^(t) was linearly decreased with age

^c^ From the model in which σ_P_^2^(t) was independent with age

^d^ For a gene, the age-specific proportion explained by the gene was calculated as the age-specific difference in σ_P_^2^(t) between the model without that gene and the model with that gene divided by the age-specific total breast cancer variance (i.e., column 2). The proportion explained by *BRCA1* and *BRCA2* was calculated as (column 2 – column 3)/column 2, the proportion explained by *BRCA1, BRCA2, PALB2, CHEK2* and *ATM* was calculated as (column 2 – column 4)/column 2, the proportion explained by *TP53* was calculated as (column 4 – column 5)/column 2, the proportion explained by the seventh hypothetical gene was calculated as (column 5 – column 6)/column 2. Where the proportion explained by a gene was negative in older ages, the proportion was assumed to be zero.

### Table S6 Families with the largest change in log-likelihood in favour of the best fitting dominant inheritance model of the hypothetical gene after fitting BRCA1, BRCA2, PALB2, CHEK2, ATM and a polygenic component

| **Family ID** | **Change in log-likelihood ^a^** | **Age at breast cancer diagnosis (years)** | | | | | **PV in other genes not considered in the best fitting dominant inheritance model of the hypothetical gene** |
| --- | --- | --- | --- | --- | --- | --- | --- |
|  |  | **Proband** | **Mother** | **Sisters** | **Aunts** | **Grandmothers** |  |
| Family 1 | 5.895 | 36 |  | 30, 36 | 27 (paternal), 28 (paternal) | 35 | *TP53* |
| Family 2 | 2.983 | 29 | 34 | 34 |  |  |  |
| Family 3 | 1.710 | 39 |  |  | 27 (paternal), 32(paternal) |  | *TP53* |
| Family 4 | 1.477 | 39 | 29 | 24 |  |  |  |
| Family 5 | 1.123 | 24 | 29 |  |  |  | *TP53* |
| Family 6 | 0.885 | 40 |  | 20, 23 |  |  |  |
| Family 7 | 0.628 | 27 | 35 |  |  |  |  |
| Family 8 | 0.497 | 29 | 34 |  |  |  |  |
| Family 9 | 0.374 | 36 |  | 33 |  |  |  |
| Family 10 | 0.328 | 41 | 36 |  | 35 |  |  |

^a^ The log-likelihood of the best fitting dominant inheritance model of the hypothetic gene minus the log-likelihood of the model including *BRCA1*, *BRCA2*, *PALB2*, *CHEK2*, *ATM* and an age-constant σ_P_^2^(t)

### Table S7 Models including BRCA1, BRCA2, PALB2, CHEK2, ATM, TP53 and a polygenic component

| **Parameter** | **Age-constant *TP53* RR ^a^** | ***TP53* RR varied by every 10 years** | ***TP53* log-RR as a linear function of age ^b^** | ***TP53* log-RR as a piecewise function of age ^c^** | ***TP53* log-RR as a linear function of age in age 20-49 years and constant in age >49 years, with an age-dependent σ_P_^2^(t) ^d^** | ***TP53* log-RR as a linear function of age in age 20-49 years and constant in age >49 years, with an age-constant σ_P_^2^(t) ^e^** |
| --- | --- | --- | --- | --- | --- | --- |
| σ_P_^2^(t) (95% CI) ^f^ | α = 2.662 (95% CI: 1.593, 3.731), β = -0.020 (95% CI: -0.039, -0.001) | α = 2.585 (95% CI: 1.424, 3.746), β = -0.019 (95% CI: -0.039, 0.002) | α = 2.600 (95% CI: 1.440, 3.761), β = -0.019 (95% CI: -0.040, 0.001) | α = 2.565 (95% CI: 1.445, 3.686), β = -0.019 (95% CI: -0.038, 0.001) | α = 2.576 (95% CI: 1.473, 3.679), β = -0.019 (95% CI: -0.038, 0.001) | 1.546 (1.389, 1.711) |
| PV frequency (95% CI) |  |  |  |  |  |  |
| *BRCA1* | 0.080%  (0.069%, 0.092%) | 0.080%  (0.069%, 0.092%) | 0.080%  (0.069%, 0.092%) | 0.080%  (0.069%, 0.092%) | 0.080%  (0.069%, 0.092%) | 0.080%  (0.069%, 0.092%) |
| *BRCA2* | 0.141%  (0.126%, 0.158%) | 0.141%  (0.126%, 0.158%) | 0.141%  (0.126%, 0.158%) | 0.141%  (0.126%, 0.158%) | 0.141%  (0.126%, 0.158%) | 0.141%  (0.126%, 0.158%) |
| *PALB2* | 0.060%  (0.049%, 0.073%) | 0.060%  (0.049%, 0.073%) | 0.060%  (0.049%, 0.073%) | 0.060%  (0.049%, 0.073%) | 0.060%  (0.049%, 0.073%) | 0.060%  (0.049%, 0.073%) |
| *CHEK2* | 0.385%  (0.338%, 0.439%) | 0.385%  (0.338%, 0.438%) | 0.385%  (0.338%, 0.438%) | 0.385%  (0.338%, 0.438%) | 0.385%  (0.338%, 0.438%) | 0.385%  (0.338%, 0.438%) |
| *ATM* | 0.167%  (0.139%, 0.200%) | 0.167%  (0.139%, 0.200%) | 0.167%  (0.139%, 0.200%) | 0.167%  (0.139%, 0.200%) | 0.167%  (0.139%, 0.200%) | 0.167%  (0.139%, 0.200%) |
| *TP53* | 0.007%  (0.005%, 0.011%) | 0.017%  (0.009%, 0.034%) | 0.018%  (0.008%, 0.041%) | 0.017%  (0.009%, 0.045%) | 0.018%  (0.009%, 0.036%) | 0.017%  (0.009%, 0.034%) |
| RR of *TP53* PV carriers (95% CI) |  |  |  |  |  |  |
| Age 20-29 years | 36.61 (19.96, 67.17) | 132.61 (55.40, 317.40) | 95.60 (41.19, 222.06) | 170.60 (70.65, 412.55) | 135.75 (61.60, 298.58) | 144.17 (66.24, 311.66) |
| Age 30-39 years | 36.61 (19.96, 67.17) | 30.26 (13.34, 68.65) | 29.83 (13.01, 67.93) | 24.03 (9.74, 59.30) | 30.59 (15.06, 62.38) | 32.13 (16.08, 64.43) |
| Age 40-49 years | 36.61 (19.96, 67.17) | 7.25 (3.07, 17.14) | 9.31 (3.89, 25.52) | 6.92 (2.40, 19.77) | 6.89 (2.48, 19.12) | 7.16 (2.66, 19.24) |
| Age 50-59 years | 36.61 (19.96, 67.17) | 3.07 (0.28, 33.04) | 2.90 (0.79, 10.81) | 2.56 (0.65, 10.11) | 2.95 (1.09, 7.95) | 3.08 (1.16, 8.17) |
| Age 60-69 years | 36.61 (19.96, 67.17) | 3.20 (0.85, 12.04) | 0.91 (0.17, 4.86) | 2.91 (0.69, 12.18) | 2.95 (1.09, 7.95) | 3.08 (1.16, 8.17) |
| Age 70-79 years | 36.61 (19.96, 67.17) | 3.55 (0, 3218679) | 0.28 (0.04, 2.24) | 4.28 (0.25, 70.11) | 2.95 (1.09, 7.95) | 3.08 (1.16, 8.17) |
| Log-likelihood | -35656.36 | 35634.98 | -35638.89 | -35635.99 | -35635.77 | -35637.37 |
| Number of parameters estimated | 9 | 14 | 10 | 15 | 11 | 10 |
| Akaike Information Criterion | 71330.72 | 71297.96 | 71297.78 | 71298.86 | 71293.54 | 71294.74 |
| P ^g^ | N/A | 4.1 × 10^-8^ | 3.4 × 10^-9^ | 7.9 × 10^-8^ | 1.1 × 10^-9^ | 7.2 × 10^-10^ |
| Best fitting model |  |  |  |  |  | Yes |

^a^ P<10^-15^ from comparing the model with its equivalent nested model including five major genes and an age-dependent polygenic component. Note that, the log-likelihood of the model including five major genes and an age-decreasing polygenic component in Table S4 is not directly comparable with the log-likelihood of the model additionally including *TP53*, because the two models included different numbers of variables. By fixing the *TP53* allele frequency to be zero and the *TP53* RR to be 1 in the model additionally including *TP53*, the model with an age-dependent σ_P_^2^(t) (essentially the same as the model including five major genes and an age-dependent σ_P_^2^(t)) had a log-likelihood of -36210.80 and an AIC of 72435.60.

^b^ Log-RR = α + β × (age – 20) in age 20-79 years, where α = 5.22 (95% CI: 4.32, 6.12), β = -0.12 (95% CI: -0.16, -0.08)

^c^ Log-RR = α + β_1_ × (age – 20) in age 20-29 years, α + 10 × β_1_ + β_2_ × (age – 30) in age 30-39 years, α + 10 × (β_1_ + β_2_) + β_3_ × (age – 40) in age 40-49 years, α + 10 × (β_1_ + β_2_ + β_3_) + β_4_ × (age – 50) in age 50-59 years, α + 10 × (β_1_ + β_2_ + β_3_ + β_4_) + β_5_ × (age – 60) in age 60-69 years, and α + 10 × (β_1_ + β_2_ + β_3_ + β_4_ + β_5_) + β_6_ × (age – 70) in age 70-79 years, where α = 6.33 (95% CI: 4.91, 7.75), β_1_ = -0.23 (95% CI: -0.40, -0.05), β_2_ = -0.17 (95% CI: -0.30, -0.04), β_3_ = -0.08 (95% CI: -0.22, 0.06), β_4_ = -0.12 (95% CI: -0.32, 0.08), β_5_ = 0.15 (95% CI: -0.12, 0.41), β_6_ = -0.07 (95% CI: -0.67, 0.53)

^d^ Log-RR = α + β × (age – 20) in age 20-49 years, where α = 5.66 (95% CI: 4.69, 6.62), β = -0.15 (95% CI: -0.21, -0.09)

^e^ Log-RR = α + β × (age – 20) in age 20-49 years, where α = 5.72 (95% CI: 4.78, 6.66), β = -0.15 (95% CI: -0.21, -0.09)

^f^ σ_P_^2^(t) = α + β × age

^g^ From the likelihood ratio test of comparing with the model including an age-constant *TP53* RR

### Table S8 Families with the largest change in log-likelihood in favour of the best fitting recessive inheritance model of the hypothetical gene after fitting BRCA1, BRCA2, PALB2, CHEK2, ATM, TP53 and a polygenic component

| **Family ID** | **Change in log likelihood ^a^** | **Age at breast cancer diagnosis (years)** | | | | |
| --- | --- | --- | --- | --- | --- | --- |
|  |  | **Proband** | **Mother** | **Sisters** | **Aunts** | **Grandmothers** |
| Family 1 | 0.676 | 29 | 34 | 34 |  |  |
| Family 2 | 0.519 | 50 | 32 | 39, 45 |  |  |
| Family 3 | 0.486 | 31 | 46 | 37 |  |  |
| Family 4 | 0.485 | 38 |  | 42, 46 | 66 (maternal) |  |
| Family 5 | 0.484 | 43 |  | 30, 30 |  |  |
| Family 6 | 0.472 | 62 | 54 | 44, 51, 53 |  | 54 (paternal) |
| Family 7 | 0.462 | 47 |  | 37, 42 |  |  |
| Family 8 | 0.437 | 45 | 46 | 38 |  | 36 (paternal) |
| Family 9 | 0.362 | 48 |  | 45, 50 |  |  |
| Family 10 | 0.329 | 46 |  | 45 |  |  |

^a^ The log-likelihood of the best fitting recessive inheritance model of the hypothetical gene minus the log-likelihood of the model including *BRCA1*, *BRCA2*, *PALB2*, *CHEK2*, *ATM*, *TP53* and an age-constant σ_P_^2^(t)

### Table S9 Sensitivity analyses results of assuming the pathogenic variant test sensitivity to be 80% for models including BRCA1, BRCA2, PALB2, CHEK2, ATM, TP53, a polygenic component, with or without a hypothetical gene

| **Parameters** | **Without the hypothetical gene** | **With the hypothetical gene** | | | |
| --- | --- | --- | --- | --- | --- |
|  | **Age-constant σ_P_^2^(t)** | **Dominant inheritance model** | **Recessive inheritance model ^a^** | **General inheritance model** | **Recessive inheritance model with an age-dependent σ_P_^2^(t) ^b^** |
| σ_P_^2^(t) (95% CI) | 1.493  (1.336, 1.658) | 1.486  (1.330, 1.652) | 1.252  (0.950, 1.595) | 1.252  (0.935, 1.615) | α = 0.742 (95% CI: -0.739, 2.223), β = 0.009 (95% CI: -0.016, 0.034) |
| PV frequency (95% CI) |  |  |  |  |  |
| *BRCA1* | 0.089%  (0.078%, 0.103%) | 0.089%  (0.077%, 0.103%) | 0.089%  (0.077%, 0.103%) | 0.089%  (0.077%, 0.103%) | 0.089%  (0.077%, 0.103%) |
| *BRCA2* | 0.157%  (0.140%, 0.177%) | 0.157%  (0.140%, 0.177%) | 0.157%  (0.140%, 0.176%) | 0.157%  (0.140%, 0.176%) | 0.157%  (0.140%, 0.176%) |
| *PALB2* | 0.067%  (0.055%, 0.082%) | 0.067%  (0.055%, 0.082%) | 0.067%  (0.055%, 0.081%) | 0.067%  (0.055%, 0.081%) | 0.067%  (0.055%, 0.081%) |
| *CHEK2* | 0.432%  (0.379%, 0.492%) | 0.432%  (0.379%, 0.492%) | 0.432%  (0.379%, 0.492%) | 0.432%  (0.379%, 0.492%) | 0.432%  (0.379%, 0.492%) |
| *ATM* | 0.187%  (0.156%, 0.224%) | 0.187%  (0.156%, 0.224%) | 0.187%  (0.156%, 0.224%) | 0.187%  (0.156%, 0.224%) | 0.187%  (0.156%, 0.224%) |
| *TP53* | 0.020%  (0.010%, 0.039%) | 0.020%  (0.008%, 0.051%) | 0.020%  (0.010%, 0.041%) | 0.020%  (0.010%, 0.042%) | 0.020%  (0.009%, 0.043%) |
| Hypothetical gene | N/A | 0.001%  (0.0001%, 0.006%) | 12.6%  (5.4%, 21.8%) | 12.6%  (5.2%, 22.5%) | 12.7%  (6.2%, 19.6%) |
| RR of *TP53* PV carriers (95% CI) |  |  |  |  |  |
| Age 20-29 years | 138.06  (64.62, 294.21) | 132.33  (54.36, 320.67) | 138.66  (60.99, 313.92) | 138.66  (59.76, 320.30) | 144.85  (59.03, 353.78) |
| Age 30-39 years | 31.13  (15.55, 62.59) | 30.30  (12.32, 74.73) | 31.32  (15.02, 65.66) | 31.32  (14.55, 67.78) | 32.41  (14.39, 73.28) |
| Age 40-49 years | 7.02  (2.58, 19.12) | 6.94  (2.05, 23.47) | 7.07  (2.57, 19.43) | 7.07  (2.50, 20.02) | 7.25  (2.52, 20.95) |
| Age 50-59 years | 2.99  (1.08, 8.30) | 2.93  (0.56, 15.37) | 3.03  (0.98, 9.37) | 3.03  (0.89, 10.33) | 3.12  (0.87, 11.21) |
| Age 60-69 years | 2.99  (1.08, 8.30) | 2.93  (0.56, 15.37) | 3.03  (0.98, 9.37) | 3.03  (0.89, 10.33) | 3.12  (0.87, 11.21) |
| Age 70-79 years | 2.99  (1.08, 8.30) | 2.93  (0.56, 15.37) | 3.03  (0.98, 9.37) | 3.03  (0.89, 10.33) | 3.12  (0.87, 11.21) |
| RR of the hypothetical gene (95% CI) |  |  |  |  |  |
| Heterozygote | N/A | 367.24  (308.71, 436.87) | 1 | 1 | 1 |
| Homozygote | N/A | 367.24  (308.71, 436.87) | 9.69  (4.09, 22.98) | 9.69  (4.06, 23.12) | 10.59  (5.01, 22.38) |
| Log-likelihood | -35636.08 | -35635.20 | -35631.95 | -35631.95 | -35631.74 |
| Number of parameters estimated | 10 | 12 | 12 | 13 | 13 |
| Akaike Information Criterion | 71292.16 | 71294.4 | 71287.90 | 71289.90 | 71289.48 |
| P ^c^ | N/A | 0.41 | 0.02 | 0.04 | 0.03 |
| Best fitting model |  |  | Yes |  |  |

^a^ Log-RR = α + β × (age – 20) in age 20-49 years, where α = 5.68 (95% CI: 4.69, 6.66), β = -0.15 (95% CI: -0.20, -0.09)

^b^ σ_P_^2^(t) = α + β × age

^c^ From the likelihood ratio test of comparing with the model including *BRCA1*, *BRCA2*, *PALB2*, *CHEK2*, *ATM*, *TP53* and an age-constant σ_P_^2^(t)

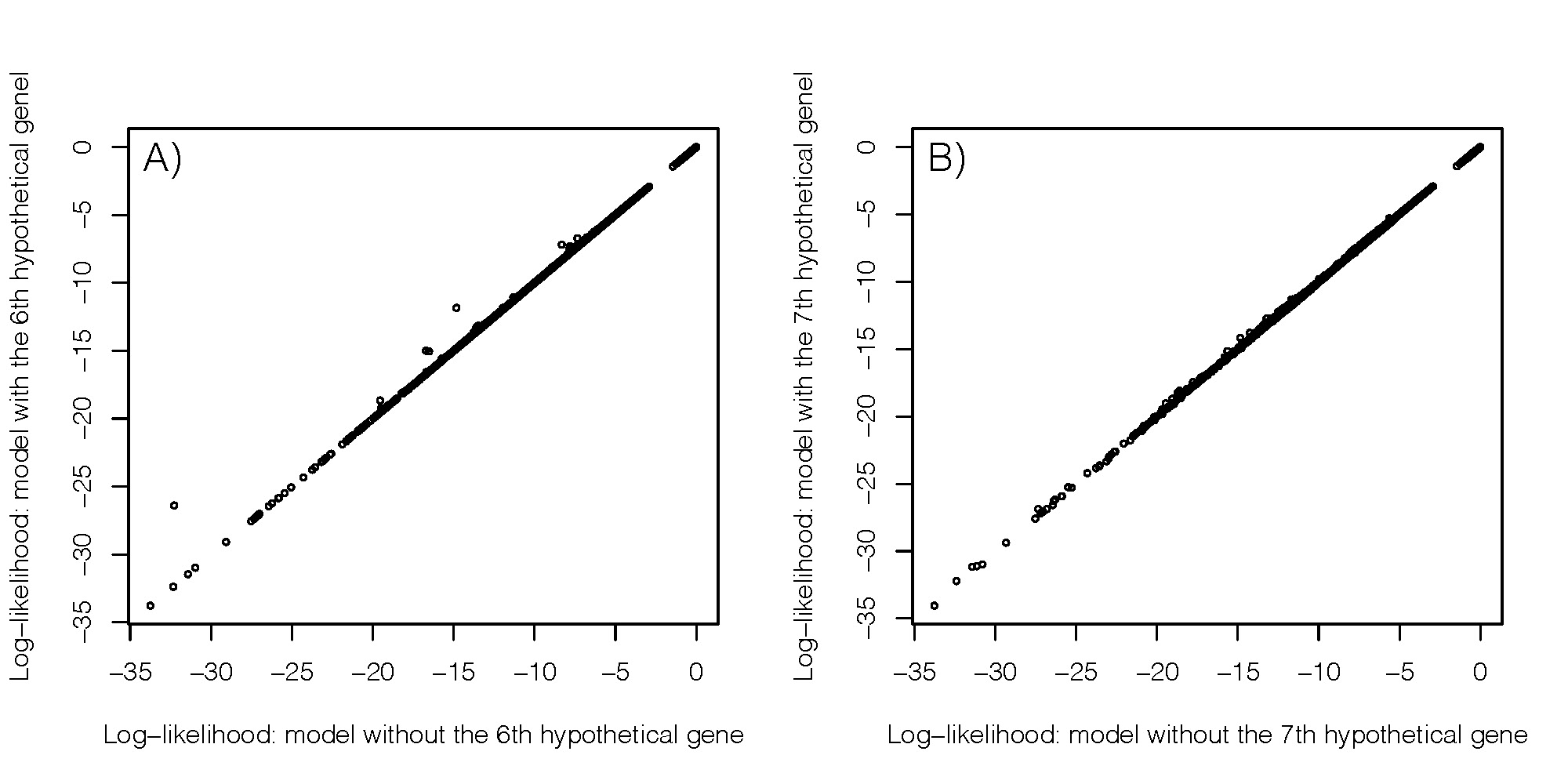

### Figure S1 Log-likelihood changes for models in favour of the sixth and seventh hypothetical gene

A) Log-likelihood changes from the analysis of the sixth hypothetical gene. X axis is the log-likelihood of the model including *BRCA1*, *BRCA2*, *PALB2*, *CHEK2*, *ATM* and an age-constant σ_P_^2^(t), and Y axis is the log-likelihood of the best fitting dominant inheritance model of the sixth hypothetical gene.

B) Log-likelihood changes from the analysis of the seventh hypothetical gene. X axis is the log-likelihood of the model including *BRCA1*, *BRCA2*, *PALB2*, *CHEK2*, *ATM*, *TP53* and an age-constant σ_P_^2^(t), and Y axis is the log-likelihood of the best fitting recessive inheritance model of the seventh hypothetical gene.
